# Distinct alterations of adiponectin, FGF-21 and IGFBP-2 link dysmetabolism with cognitive decline across the Alzheimer’s disease spectrum

**DOI:** 10.1101/2025.11.26.25340689

**Authors:** C. Dallaire-Théroux, H.L. Denis, R. Cottez, C. Tremblay, A. Provencher, J. Valentin-Escalera, M. Leclerc, A. Loiselle, M. Tournissac, O. Potvin, S. Belleville, A. Gangloff, F. Picard, H. Zetterberg, Consortium for the Early Identification of Alzheimer’s Disease – Quebec (CIMA-Q), F. Calon

**Affiliations:** Neurosciences Axis, Centre de recherche du CHU de Québec - Université Laval, Quebec City, QC, Canada; Division of Neuroscience, Hôpital de l’Enfant-Jésus, Centre Hospitalier Universitaire (CHU) de Québec-Université Laval, Quebec City, QC, Canada; Faculty of Pharmacy, Université Laval, Quebec City, QC, Canada; Quebec Heart and Lung Research Institute (IUCPQ), Quebec City, QC, Canada; Research centre, Institut universitaire de gériatrie de Montréal, Montreal, QC, Canada; Oncology research axis, Centre de recherche du CHU de Québec - Université Laval, Quebec City, QC, Canada; Department of Psychiatry and Neurochemistry, Institute of Neuroscience and Physiology, Sahlgrenska Academy of the University of Gothenburg, Gothenburg, Sweden; Clinical Neurochemistry Laboratory, Sahlgrenska University Hospital, Mölndal, Sweden; Department of Pathology and Laboratory Medicine, University of Wisconsin School of Medicine and Public Health, Madison, WI, USA; Wisconsin Alzheimer’s Disease Research Center, University of Wisconsin School of Medicine and Public Health, University of Wisconsin-Madison, Madison, WI, USA; Department of Neurodegenerative Disease, UCL Institute of Neurology, Queen Square, London, UK; UK Dementia Research Institute at UCL, London, UK; Centre for Brain Research, Indian Institute of Science, Bangalore, India

**Keywords:** Alzheimer’s Disease, adiponectin, FGF-21, IGFBP-2, metabolic syndrome, metabolism, mild cognitive impairment, subjective cognitive decline, Consortium for the early identification of Alzheimer’s Disease – Quebec, CIMA-Q

## Abstract

**Introduction:** Metabolic disorders are risk factors for Alzheimer’s disease (AD), although underlying mechanisms remain unclear. We investigated the relationship between peripheral metabolic markers – adiponectin, FGF-21 and IGFBP-2 – and AD.

**Methods:** Participants with subjective cognitive decline (SCD), mild cognitive impairment (MCI), AD and cognitively healthy controls (CH) were from the CIMA-Q cohort (n=287). Serum adiponectin, FGF-21, and IGFBP-2 concentrations were measured, compared between groups, and assessed for associations with clinical, cognitive, biochemical and MRI data.

**Results:** Metabolic dysfunction was linked to lower adiponectin and IGFBP-2, but higher FGF-21. Both FGF-21 and IGFBP-2 increased with age and were inversely associated with cognitive performance. IGFBP-2 was elevated at SCD stage and correlated with plasma pTau181 and amygdala atrophy. Adiponectin was unrelated to cognition.

**Discussion:** These findings suggest that IGFBP-2, and FGF-21 to a lesser extent, may serve as early biomarkers of cognitive impairment, reflecting intricate links between peripheral dysmetabolism and AD.

## 1. BACKGROUND

The association between metabolic disorders and Alzheimer’s disease (AD) is now very well established. It has been estimated that patients with metabolic syndrome are over three times more likely to develop AD [1, 2]. Evidence from numerous epidemiological studies reveals that mid-life obesity, type-2 diabetes and high blood pressure are potentially modifiable risk factors for late-life dementia and AD [3, 4]. Metabolic dysfunctions in AD are therefore being investigated for their possible early involvement in pathophysiology and for the development of new treatments [5]. Although the links between metabolic diseases, insulin resistance (peripheral or central) and cognitive decline has been confirmed by several studies [1, 6, 7], they do not appear to be directly explained by AD neuropathology [8], underscoring the importance of peripheral mechanisms underlying these associations.

Among all peripheral markers related to the insulin signaling pathway, serum adiponectin, fibroblast growth factor 21 (FGF-21) and insulin-like growth factor-binding protein 2 (IGFBP-2) were identified as three promising candidates. Adiponectin is a protein predominantly secreted by mature adipocytes and is thought to be involved in a number of metabolic processes, such as the regulation of glucose homeostasis, fatty acid oxidation in muscles and insulin resistance [9]. FGF-21 is a metabolic hormone synthesized by various tissues including adipose tissue (refers to adipokines), liver (refers to hepatokines), and muscle (refers to myokines). A recent study suggest that blood FGF-21/adiponectin ratio may predict the onset of prediabetes, diabetes and might be used as a biomarker of deterioration in glycemia [10]. FGF-21 is also involved in glucose homeostasis regulation and provide efficient and durable glucose control and triglyceride lowering in diabetic animals [11]. IGFBP-2 is a protein mainly produced in the liver that binds to insulin-like growth factor (IGF)-1 and IGF-2, thereby regulating IGF bioavailability, while also engaging in mechanisms independent of IGF signaling [12, 13]. Low circulating IGFBP-2 concentrations have been linked to dyslipidemia, insulin resistance and metabolic syndrome [14, 15]. IGFBP-2 is most particularly found in the central nervous system and is involved in brain development as well as in regulation of neuronal plasticity [16]. However, despite being altered in dysmetabolism, these three metabolic determinants can have very different mechanistic roles, and their impact on brain function remains to be determined.

Over the last years, adiponectin [17–21], FGF-21 [22, 23] and IGFBP-2 [24–35], have been investigated in animal models and subjects with cognitive impairment and dementia attributed to AD. However, while most studies reported that higher levels of IGFBP-2 levels in the blood and CSF are associated with AD (including cognition, magnetic resonance imaging (MRI) and cerebrospinal fluid (CSF) tau) [24–26, 28, 30, 31, 34, 35], the pattern of metabolic hormone changes in the early stages of AD remains poorly understood due to a lack of studies involving clearly defined clinical cohorts.

Here we aimed to explore the relationship between peripheral markers of metabolic function, including serum adiponectin, FGF-21 and IGFBP-2, with clinical outcomes in the AD continuum. More specifically, we investigated their associations with various metabolic parameters (body mass index (BMI), markers of glycemic control and lipid profile, among others), cognitive performance (both global and domain-specific), AD fluid-based biomarkers and MRI metrics using blood samples and clinical data from the Consortium for early identification of Alzheimer’s disease-Quebec (CIMA-Q) cohort [36]. The CIMA-Q cohort provides comprehensive, well-characterized longitudinal data encompassing clinical, cognitive, biological, and neuroimaging measures, making it an invaluable resource for exploring early biomarkers and underlying mechanisms in AD. Its standardized protocols and emphasis on early-stage participants further enhance the robustness and translational value of the findings. Through this approach, we sought to determine how peripheral markers of metabolic function evolve in the early stages of AD to better understand their role in the progression of the disease and to establish their potential as biomarkers of cognitive decline.

## 2. METHODS

### 2.1 Participants

The study sample includes older adults recruited from the CIMA-Q cohort. The main objective of the CIMA-Q is to characterize and track AD progression in individuals at higher risk of dementia, such as those presenting with subjective cognitive complaints, and to propose valuable data for identification of AD biomarkers. The recruitment protocol and all clinical, cognitive, and neuropsychiatric data collection procedures have been previously described [36]. Briefly, in its first phase starting in 2013, CIMA-Q recruited 290 participants aged 65 years and above who were fluent in French or English and who lived in the province of Quebec, Canada. The participants were characterized on physical, clinical, and cognitive levels through extensive evaluations. They could consent to blood and CSF collection, MRI, positron emission tomography (PET), cryopreservation of peripheral blood mononuclear cells, and post-mortem brain retrieval.

Data for the present study were obtained at baseline, or first visit, from 287 CIMA-Q participants from whom serum was sampled. These participants were classified into four groups according to clinical diagnoses provided by expert physicians: cognitively healthy (CH; n=56), subjective cognitive decline (SCD; n=115), mild cognitive impairment (MCI; n=85) and AD (n=31) (**Table 1**). CH participants had no cognitive impairment nor complaints concerning their memory. Identification of SCD was based on the Subjective Cognitive Decline Initiative research criteria, clinically defined as a self-experienced persistent decline in cognitive capacity compared to a previously normal state, without any objective cognitive impairment on standardized cognitive testing, and not explained by an acute event or other medical condition [40]. MCI and AD diagnoses were based on the National Institute on Aging and Alzheimer’s Association criteria [36, 37]. Inclusion and exclusion criteria for each group can be found in **Supplemental Material Appendix A**.

**Table 1.** Participant Characteristics Including Clinical, Sociodemographic and Blood Assessment Data in Each Clinical Diagnosis Group.

|  |  |  |  |  | *P* values |  |  |  |  |  |  |
| --- | --- | --- | --- | --- | --- | --- | --- | --- | --- | --- | --- |
|  | CH<br>N=56 | SCD<br>N=115 | MCI<br>N=85 | AD<br>N=31 |  | CH vs.<br>SCD | CH vs.<br>MCI | CH vs.<br>AD | SCD vs.<br>MCI | SCD<br>vs. AD | MCI<br>vs. AD |
| Participant Characteristics |  |  |  |  |  |  |  |  |  |  |  |
| Age(years) | 73.5(6) | 72.5(5) | 76.4(6) | 77.5(7) | **<.001** | >.99 | **.015** | **.031** | **<.001** | **<.001** | >.99 |
| Sex(M/F) | 17/39 | 32/83 | 42/43 | 15/16 | **.006** | .73 | **.025** | .10 | **.002** | **.030** | .92 |
| APOE ε4 carrier(%) | 16 | 24 | 33 | 48 | **.005** | .27 | **.026** | **.001** | .14 | **.007** | .13 |
| Clinical and Sociodemographic Data |  |  |  |  |  |  |  |  |  |  |  |
| MNA(/14) | 13(1) | 13(2) | 13(2) | 13(1) | .88 | >.99 | >.99 | >.99 | >.99 | >.99 | >.99 |
| Education(years)* | 15(4) | 15(4) | 14(4) | 16(5) | .09 | >.99 | >.99 | .81 | .81 | .78 | .08 |
| Waist size(cm) | 97(12) | 96(13) | 98(11) | 96(12) | .72 | >.99 | >.99 | >.99 | >.99 | >.99 | >.99 |
| Weight(kg) | 69(13) | 69(16) | 73(12) | 71(13) | .10 | >.99 | .41 | >.99 | .13 | >.99 | >.99 |
| BMI(kg/m<sup>2</sup>) | 26(4) | 27(5) | 27(4) | 26(4) | .42 | >.99 | .699 | >.99 | >.99 | >.99 | >.99 |
| Diastolic(mm Hg) | 76(8) | 76(8) | 76(8) | 75(8) | .52 | .98 | .98 | .92 | >.99 | .72 | .75 |
| Systolic(mm Hg) | 133(17) | 136(18) | 138(20) | 139(21) | .51 | >.99 | >.99 | >.99 | >.99 | >.99 | >.99 |
| Blood Assessment |  |  |  |  |  |  |  |  |  |  |  |
| Blood cells |  |  |  |  |  |  |  |  |  |  |  |
| Leukocytes(x10<sup>9</sup>/L) | 5.5(1.3) | 5.6(1.5) | 6.2(2.0) | 5.9(1.4) | .22 | >.99 | .78 | >.99 | .40 | >.99 | >.99 |
| Erythrocytes(x10<sup>12</sup>/L) | 4.5(.4) | 4.6(.4) | 4.6(.4) | 4.7(.4) | .32 | >.99 | >.99 | .41 | >.99 | .80 | >.99 |
| Hemoglobin(g/L) | 138(11) | 139(10) | 141(10) | 142(13) | .35 | >.99 | .71 | .93 | >.99 | >.99 | >.99 |
| Platelets(x10<sup>9</sup>/L) | 215(57) | 237(47) | 220(48) | 209(43) | .05 | .28 | >.99 | >.99 | .18 | .22 | >.99 |
| Glycemia index |  |  |  |  |  |  |  |  |  |  |  |
| Glycemia(mmol/L) | 5.7(1.0) | 5.5(1.0) | 5.6(1.2) | 5.7(.9) | .35 | >.99 | >.99 | >.99 | >.99 | .63 | >.99 |
| HbA1c(%) | 5.7(.7) | 5.5(.6) | 5.7(.8) | 5.7(.4) | .11 | >.99 | >.99 | >.99 | .48 | .16 | >.99 |
| Vitamins & Hormone |  |  |  |  |  |  |  |  |  |  |  |
| Vitamin B<sub>12</sub>(pmol/L) | 254(99) | 308(158) | 318(198) | 297(161) | .22 | .23 | >.99 | >.99 | >.99 | >.99 | >.99 |
| Vitamin B<sub>9</sub>(mmol/L) | 38(10) | 39(11) | 37(11) | 27(10) | .21 | >.99 | >.99 | .42 | >.99 | .23 | .66 |
| TSH(mU/L) | 2.7(1.3) | 3.1(1.7) | 2.7(1.7) | 3.1(1.9) | .39 | >.99 | >.99 | >.99 | .576 | >.99 | >.99 |
| Inflammation |  |  |  |  |  |  |  |  |  |  |  |
| C-reactive protein(mg/L) | 2.7(2.5) | 2.3(2.9) | 2.6(2.7) | 3.2(4.6) | .68 | >.99 | >.99 | >.99 | >.99 | >.99 | >.99 |
| Renal function |  |  |  |  |  |  |  |  |  |  |  |
| Blood Urea Nitrogen(mmol/L) | 5.9(1.8) | 5.5(1.6) | 5.7(1.5) | 7.5(2.4) | **.0003** | >.99 | >.99 | **.012** | >.99 | **<.001** | **.001** |
| Creatinine(μmol/L) | 81(22) | 77(19) | 83(18) | 101(38) | **.0014** | >.99 | >.99 | **.020** | .19 | **.0012** | .163 |
| DFG(mL/min/1.73m<sup>2</sup>) | 75(10) | 75(14) | 70(14) | 78(11) | .29 | >.99 | >.99 | >.99 | .63 | >.99 | .67 |
| Hepatic function |  |  |  |  |  |  |  |  |  |  |  |
| ALT | 20(9) | 21(8) | 22(10) | 19(5) | .62 | >.99 | >.99 | >.99 | >.99 | >.99 | >.99 |
| AST | 23(7) | 23(5) | 24(7) | 23(4) | .64 | >.99 | >.99 | >.99 | >.99 | >.99 | >.99 |
| AST/ALT | .9(.2) | .9(.2) | .9(.3) | .9(.3) | .23 | .71 | >.99 | >.99 | >.99 | .57 | >.99 |
| LAP(IU/L) | 64(20) | 62(19) | 64(18) | 66(16) | .49 | >.99 | >.99 | >.99 | >.99 | >.99 | >.99 |
| Lipidic profile |  |  |  |  |  |  |  |  |  |  |  |
| TG(mmol/L) | 1.2(.6) | 1.3(.7) | 1.4(.7) | 1.2(.4) | .67 | >.99 | >.99 | >.99 | >.99 | >.99 | >.99 |
| Cholesterol(mmol/L) | 5.1(1.1) | 5.3(1.2) | 4.9(1.0) | 5.1(1.5) | .34 | >.99 | >.99 | >.99 | .53 | >.99 | >.99 |
| HDL(mmol/L) | 1.5(.5) | 1.6(.5) | 1.5(.4) | 1.6(.4) | .35 | >.99 | >.99 | >.99 | .70 | >.99 | >.99 |
| Cholesterol/HDL | 3.5(.9) | 3.4(.9) | 3.5(.9) | 3.2(.9) | .38 | >.99 | >.99 | >.99 | >.99 | >.99 | >.99 |
| LDL(mmol/L) | 3.1(.9) | 3.2(1.0) | 2.8(.8) | 3.0(1.2) | .30 | >.99 | >.99 | >.99 | .43 | >.99 | >.99 |

Statistical analyses: Values are means(SD). Normality test used Agostino & Pearson test. When data was normally distributed, a one-way ANOVA test followed by Tukey's post-hoc tests was performed. When data were not normally distributed, a nonparametric Kruskal-Wallis test followed by Dunn's post-hoc test was performed. A Chi Square test of independence was performed for sex and *APOE* ε4 genotype. Bold values indicate $p<0.05$ . \*For education, data was not available for 7 CH, 13 SCD, 10 MCI, and 1 AD participants. However, the information about the highest level of education completed (primary, high school, college, or university) was obtained for all cases. **Abbreviations:** AD, Alzheimer's disease; Alp, alkaline phosphatase; ALT, alanine transaminase; AST, aspartate; CH, cognitively healthy; DFG, Glomerular Filtration Rate; HbA1c, Glycated hemoglobin; HDL, high-density lipoprotein; LAP, leukocyte alkaline phosphatase; LDL, low-density lipoprotein; MCI, mild cognitive impairment; MNA, Mini Nutritional Assessment; SCD, subjective cognitive decline; TG, triglycerides; TSH, thyroid stimulating hormone.

The CIMA-Q study protocol and the present study were both approved by the Aging-Neuroimaging Research Ethics Committee of the CIUSSS du Centre-Sud-de-Île-de-Montréal (CER VN 14-15-19 and CER VN 18-19-01) and the Research Ethics Committee of the CHU du Québec Research Center (2015-2366, 2019-4458, 2019-4460 and 2022-6290). The study was conducted in accordance with the ethical principles outlined in the 1964 Declaration of Helsinki and its subsequent amendments. All participants provided informed written consent.

### 2.2 Blood collection and processing

Blood samples were obtained in early morning from each participant under fasting conditions (minimum of eight hours). Each sample was mixed by inverting the tube and then kept upright at room temperature before being processed. Blood samples used for serum extraction were placed directly on ice for 30 minutes to allow coagulation. The SST tubes were inverted five times and centrifuged 4°C for fifteen minutes at 1300 x g. Serums where then aliquoted and stored at -80°C for long-term storage. A routine blood analysis was performed as detailed in **Supplemental Material Appendix B**. Serum levels of adiponectin, FGF-21 and IGFBP-2 were obtained using enzyme-linked immunosorbent assays (ELISA) following manufacturer’s instructions (Human Total Adiponectin/Acrp30 Quantikine ELISA Kit, #DFRP300; Human FGF-21 Quantikine ELISA Kit, #DF2100; Human IGFBP-2 Quantikine ELISA Kit, #DGB200, all from R&D Systems, USA). For pTau-217, the Meso Scale Discovery technology was used (S-PLEX Human pTau217 Kit, #K151APFS, MSD, USA). Both pTau181 and pTau231 were assessed with Simoa technology [38, 39]. Although plasma amyloid beta (Aβ) was not measured at baseline in the CIMA-Q cohort, evidence indicates that plasma pTau217 is a robust marker reflecting both the presence and extent of cerebral Aβ burden in AD [40–42]. *APOE* genotypes are shown in **Table 1**. More methodological details can be found in our previous publication [36].

### 2.3 CSF collection and processing

Lumbar punctures were performed by an anesthesiologist or a neurologist on the participants who consented to the procedure. Between 10 and 15 ml of CSF were collected under non-fasting conditions. Within 15 minutes of the puncture, the sample was gently mixed by inverting the tube and centrifuging it at room temperature for ten minutes at 2000 x g using a Sorvall RT 6000D centrifuge. Next, the CSF was aliquoted into 0.1 ml fractions using low protein-binding polypropylene microtubes which had previously been kept on dry ice for an hour. Finally, the aliquots were placed on dry ice within 60 minutes of the puncture and placed in long-term storage at -80°C. Amyloid beta (Aβ) peptides levels of 38, 40, and 42 amino acids and total tau protein levels were obtained using a multiplexed immunoassay with electrochemiluminescence detection following manufacturer’s instructions (Aβ Peptide Panel 1 Kit (6E10), #K15200E and Human Total Tau Kit, #K151LAE-1, Meso Scale Discovery, Rockville, MD, USA). More details can be found in our previous study [36].

### 2.4 Magnetic resonance imaging acquisition and processing

Participants who consented and who were eligible underwent MRI at a magnetic field strength of 3 Tesla at five sites with either Siemens Healthcare (TrioTim and Prisma Fit) or Philips Medical Systems (Achieva and Ingenia) scanners. The acquisitions followed the Canadian Dementia Imaging Protocol (https://www.cdip-pcid.ca): T1-weighted, proton density, T2-weighted, fluid-attenuated inversion recovery (FLAIR), T2-star–weighted sequences. All scans were first visually inspected by a radiologic technologist blinded to the diagnosis. We based our analyses on the 163 participants from CIMA-Q study who underwent MRI (CH, n=35; SCD, n=79; MCI, n=37; AD, n=12). Morphometry measurements were derived using *FreeSurfer* 6.0 (http://freesurfer.net) using fully automated directive parameters (no manual intervention or expert flag options). Brain segmentations were visually inspected by O.P. through at least 20 evenly distributed axial and coronal section. For each region, we verified that segmentation did not have significant inadequate inclusion (e.g. dura mater, ventricle) or omission of approximately 100 voxels or more. Following inspection, no failed segmentation was detected. Then, using the NOMIS tool, morphometric measures were transformed into adjusted Z scores (referred to as standardized brain volumes) for estimated intra-cranial volume and image quality and resolution. Our volumetric regions of interest were the whole brain, cortex, white and gray matter, ventricles, and hippocampus and amygdala. References to protocols and tools can be found in **Supplemental Material Appendix C.**

### 2.5 Cognitive and behavioral assessments

The following tests were performed to obtain a detailed phenotypical characterization of participants and then correlate it with biological findings. Cognitive functioning was measured by the Montreal Cognitive Assessment (MoCA), the Telephone Mini-Mental State examination (tMMSE), the Face-Name Association (FNAME), the Boston Naming Test (BNT), the Stroop version (card 3) of the Delis-Kaplan Executive Function System, parts A and B of the Trail Making Test (TMT), the Logical Memory Task from the Wechsler Memory Scale, the WAIS Digit Symbol Substitution Test (WDSST), and the Rey Auditory Verbal Learning Test (RAVLT). Mental health was assessed using the Geriatric Depression Scale GDS), the Patient Health Questionnaire (PHQ) and the Geriatric Anxiety Inventory (GAI). References to neuropsychological tests can be found in **Supplemental Material Appendix C.**

### 2.6 Statistical analysis

For group comparison in **Figures 1** and **6**, nonparametric Kruskal-Wallis tests were used to compare groups, followed by Dunn’s post-hoc test (**Figure 1A,E,I**; **Figure 6A**). Alternatively, an ANCOVA was performed after adjustment for age and sex followed by Dunnett’s post-hoc test to compare metabolic analytes between clinical groups (**Figure 1B,F,J**; **Figure 6B**). Spearman correlations were used to assess the relationships between adiponectin, FGF-21, IGFBP-2 and age (**Figure 1C,G,K**), clinical and sociodemographic data, blood assessment (**Figure 2**), clinical scores (**Figure 3**), AD biomarkers (**Figure 4**) and brain volumes (**Figure 5**). Spearman correlation p-value of the adjusted and non-adjusted metabolic analytes has been reported in figures (**Figure 1C,G, K**; **Figure 2-5**). In Figures representing pairwise comparisons, data are expressed as means ± standard error of the mean (SEM). Missing data were handled using an available-case analysis (or pairwise deletion), whereby each analysis included all participants with complete data for the variables involved. Results were considered statistically significant when *p* < 0.05. All statistical analyses mentioned above were performed using GraphPad Prism 10.5.0 (GraphPad Software Inc., San Diego, CA, USA), JMP Pro 18.2.2 (JMP Statistical Discovery LLC, Cary, NC, USA), or RStudio 2024.12.1+563 (https://www.r-project.org/) software.

**Figure 1.**
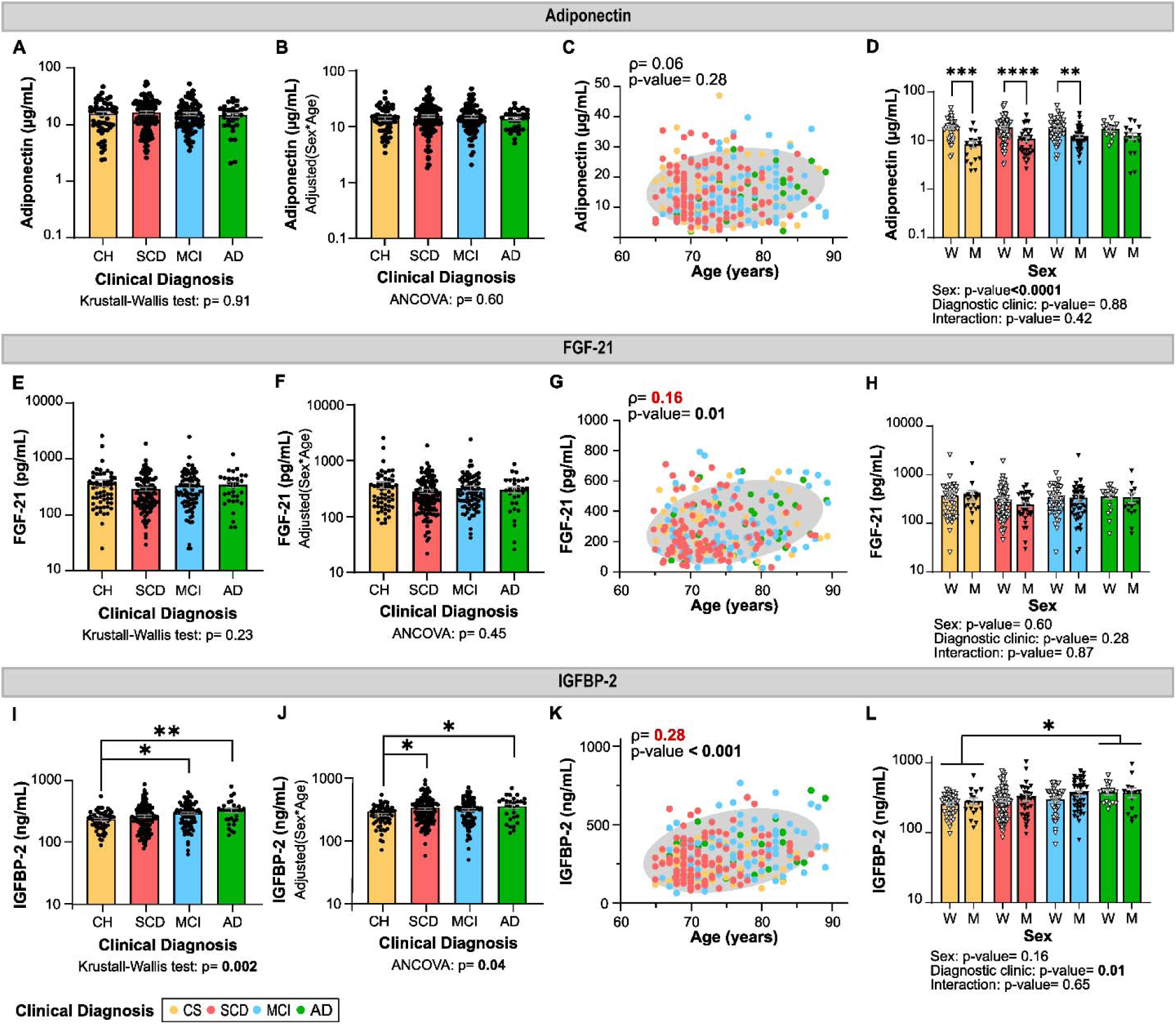
Interaction Between Clinical Diagnosis Group, Age, Sex and Serum Adiponectin, FGF-21 and IGFBP-2. Association between serum levels of adiponectin, FGF-21 and IGFBP-2 and clinical diagnosis before (respectively **A,E,I**) and after adjustment for age and sex (respectively **B,F,J**). Correlation between adiponectin, FGF-21 and IGFBP-2 and age (respectively **C,G,K**). Serum levels of metabolites separated by clinical diagnoses and by sex (respectively **D,H,L**). Statistical analyses: Values are means ± SEM. Nonparametric Kruskal-Wallis tests followed by Dunn’s post-hoc test were performed for group comparisons (A, E I). ANCOVA, adjusted for age and sex, were conducted, followed by Dunnett’s post-hoc tests for pairwise comparisons (B, F, J). Linear regression between metabolic hormones and age are presented with rho (ρ) and p-values (Spearman) (C, G, K). Two-way ANOVA was performed using clinical diagnosis and sex as variables (D, H, L). *p<0.05; **p<0.01; ***p<0.005; ****p<0.001. Abbreviations: AD, Alzheimer’s disease; CH, cognitively healthy; MCI, mild cognitive impairment; SCD, subjective cognitive decline; SEM, standard error of the mean.

**Figure 2.**
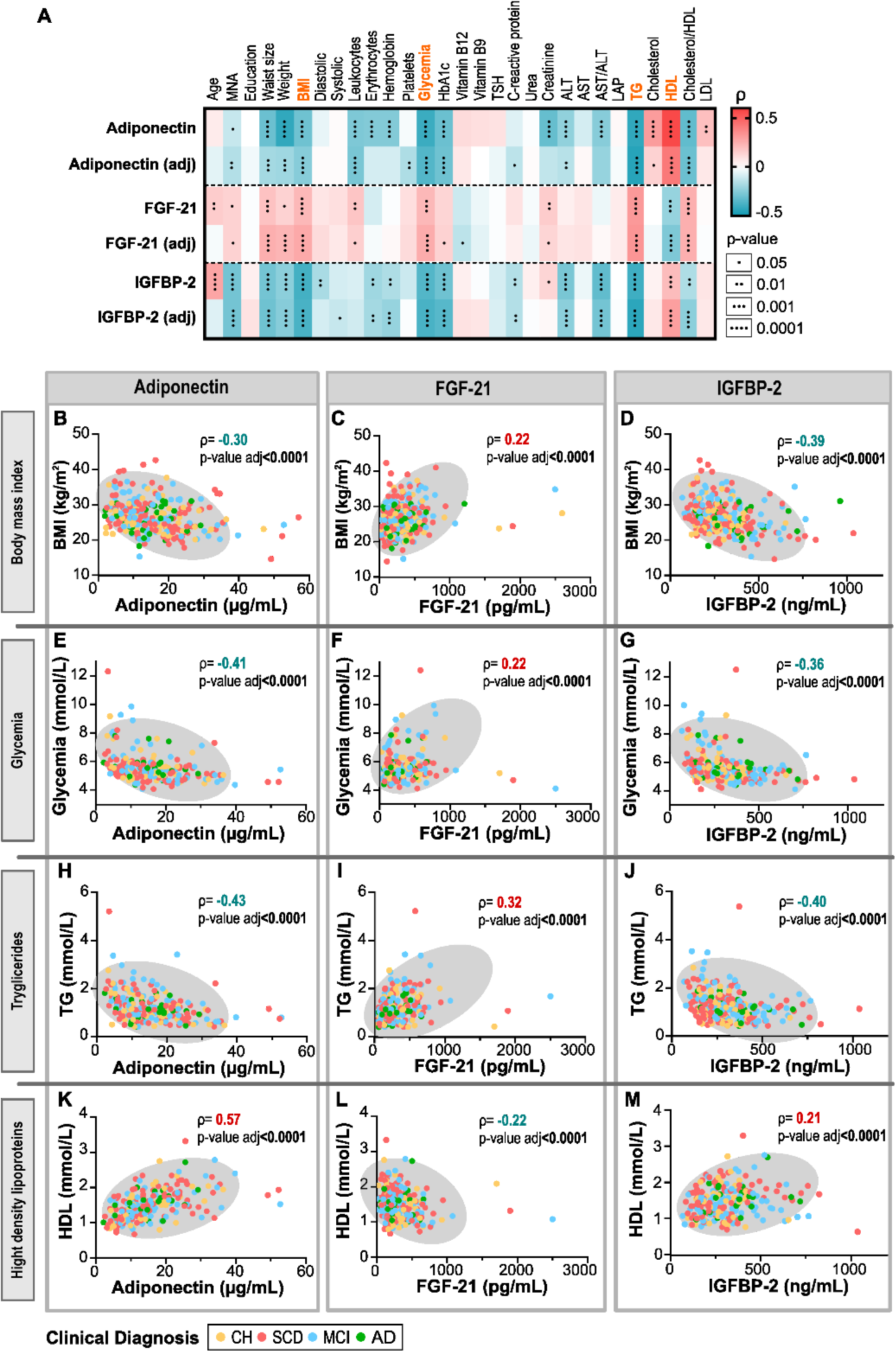
Multiple Linear Regression for Adiponectin, FGF-21 and IGFBP-2 on Clinical and Sociodemographic Data & Blood Assessment. Statistical analyses: Red and blue highlighted cells respectively indicate positive and negative correlations. Both unadjusted and adjusted p-values (for age and sex) are presented from Spearman correlation analyses (A). Linear regression between metabolic hormones and selected markers of metabolic status (highlighted in orange in A) are presented with rho (ρ) and p-values (Spearman) (B-M). Abbreviations: AD, Alzheimer’s disease; adj, adjusted for age and sex; Alp, alkaline phosphatase; ALT, alanine transaminase; AST, aspartate; CH, cognitively healthy; HbA1c, Glycated hemoglobin; HDL, high-density lipoprotein; LAP, leukocyte alkaline phosphatase; LDL, low-density lipoprotein; MCI, mild cognitive impairment; MNA, Mini Nutritional Assessment; SCD, subjective cognitive decline; TG, triglycerides; TSH, thyroid stimulating hormone.

**Figure 3.**
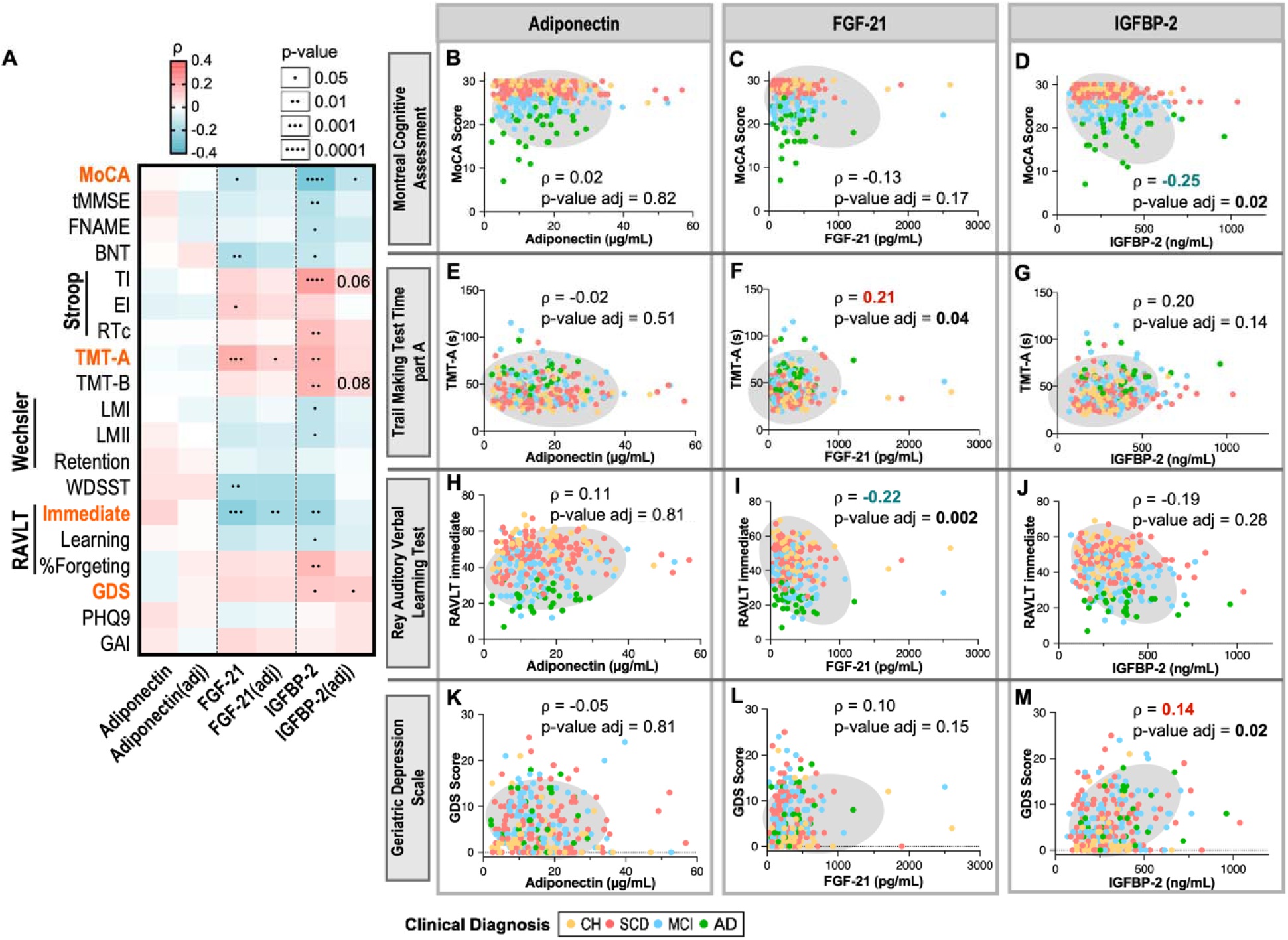
Association Between Serum Adiponectin, FGF-21, IGFBP-2 and Cognitive Scores. Statistical analyses: Red and blue highlighted cells respectively indicate positive and negative correlations. Both unadjusted and adjusted p-values (for age and sex) from Spearman correlation analyses are shown. (A). Linear regression between metabolic hormones and selected scores in neuropsychology tests and clinical scales (highlighted in orange in A) are presented with rho (ρ) and p-values (Spearman) (B-J). Abbreviations: AD, Alzheimer’s disease; adj, adjusted for age and sex; BNT, Boston Naming Test - correct spontaneous answers; CH, cognitively healthy; CSF, cerebrospinal fluid; EI score, error interference score; FNAME, Face-Name Associative Memory Exam; GAI, Geriatric Anxiety Inventory; GDS, Geriatric Depression Scale; LMI, Logical Memory task, immediate recall from the Wechsler Memory Scale; LMII, Logical Memory task, delayed recall from the Wechsler Memory Scale; MCI, mild cognitive impairment; MNA, Mini Nutritional Assessment; MoCA, Montreal Cognitive Assessment; PHQ9, Patient Health Questionnaire; RAVLT, Rey Auditory Verbal Learning Test; RTc, Reaction Time corrected (Card 3); SCD, subjective cognitive decline; TI score, time interference score; T-MMSE, Telephone Mini-Mental State Examination; TMT, Trail Making test Time; WDSST, WAIS Digit Symbol Substitution Test.

**Figure 4.**
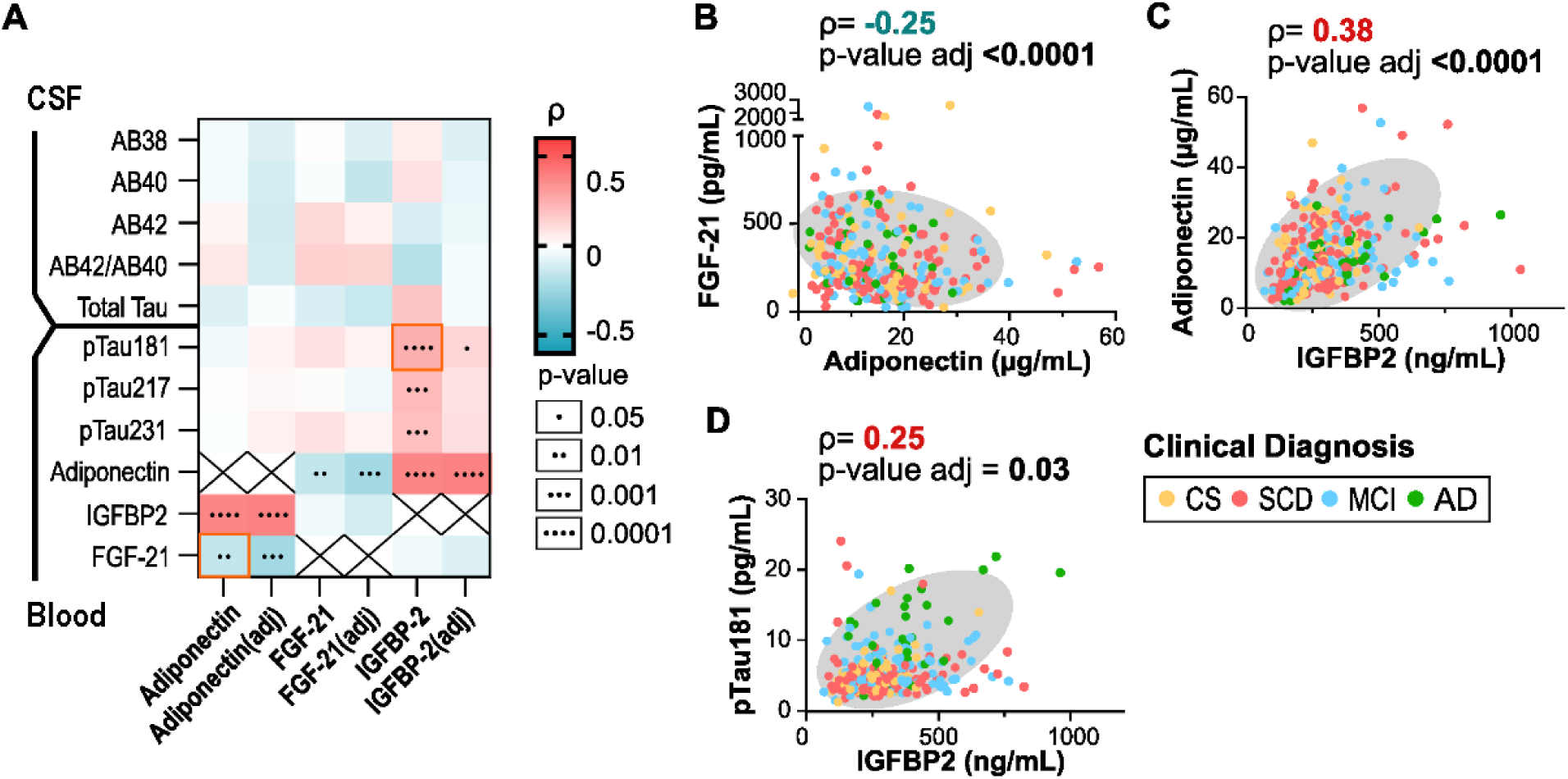
Associations Between Adiponectin, FGF-21 and IGFBP-2 and AD Biomarkers. Statistical analyses: Red and blue highlighted cells respectively indicate positive and negative correlations. Both unadjusted and adjusted p-values (for age and sex) from Spearman correlation analyses are shown. (A). Selected linear regression are presented with rho (ρ) and p-values (Spearman) (B-D). Abbreviations: AD, Alzheimer’s disease; adj, adjusted for age and sex; CH, cognitively healthy; MCI, mild cognitive impairment; SCD, subjective cognitive decline.

**Figure 5.**
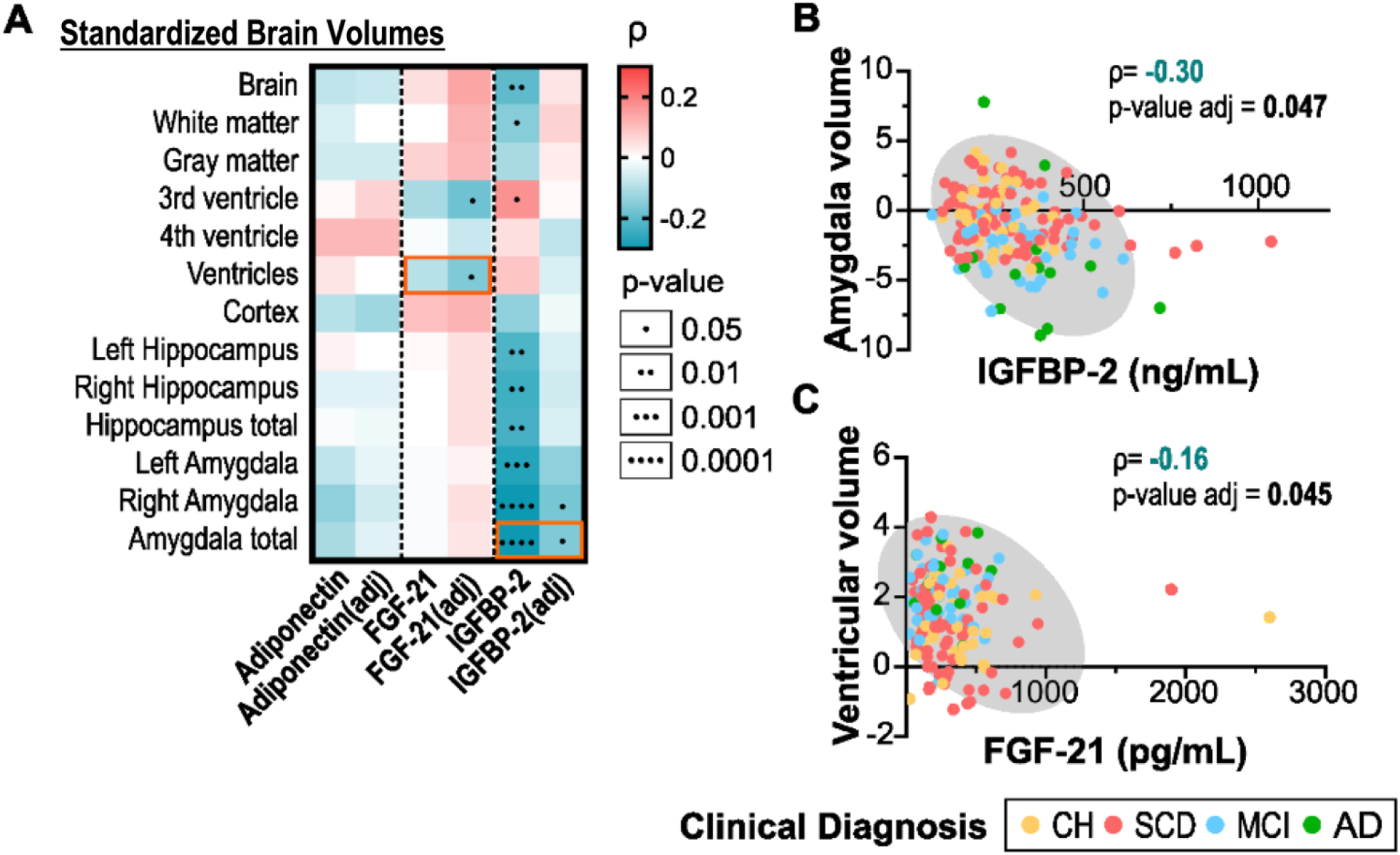
Multiple Linear Regression Modeling of Standardized Brain Volumes in Relation to Adiponectin, FGF-21, and IGFBP-2. Statistical analyses: Red and blue highlighted cells respectively indicate positive and negative correlations. Both unadjusted and adjusted p-values (for age and sex) from Spearman correlation analyses are shown. (A). Selected linear regression between metabolic hormones and MRI metrics are presented with rho (ρ) and p-values (Spearman) (B-J). Abbreviations: AD, Alzheimer’s disease; adj, adjusted for age and sex; CH, cognitively healthy; MCI, mild cognitive impairment; SCD, subjective cognitive decline.

**Figure 6.**
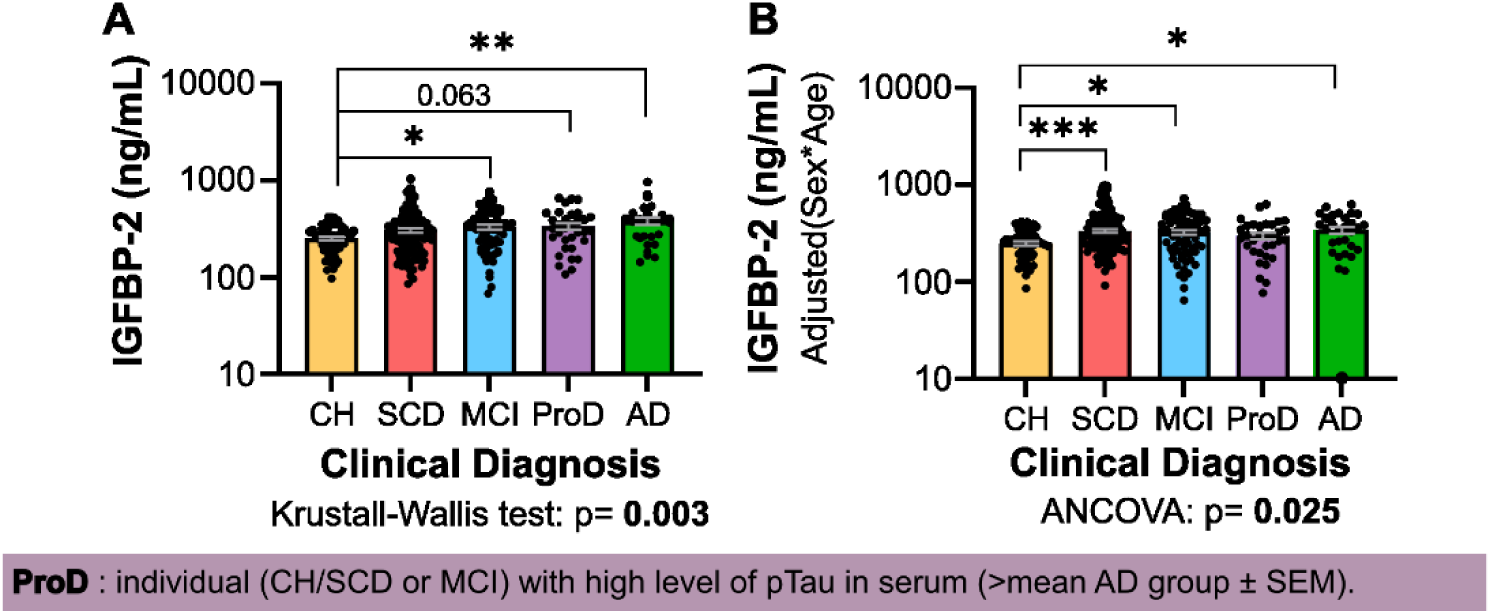
Interaction Between IGFBP-2 Levels and Prodomal. Association between clinical diagnosis and serum levels of IGFBP-2 (**A**) and after adjustment with age and sex (**B**). The threshold for a biological or prodromal diagnosis was set at 8 pg/ml, which corresponds to one standard error of the mean lower than the mean of the three tau biomarkers (pTau181, pTau217, and pTau231) in subjects with an AD diagnosis. Individuals who had a value of pTau181, pTau217, or pTau231 above 8 pg/ml were considered prodromal. Statistical analyses: Values are means (SEM) shown a logarithmic scale. A nonparametric Kruskal-Wallis test followed by Dunn’s post-hoc test was performed for group comparisons (A). An ANCOVA, adjusted for age and sex, was conducted, followed by Dunnett’s post-hoc test (B). *p<0.05; **p<0.01; $p<0.05 vs CS group. Abbreviations: AD, Alzheimer’s disease; CH, cognitively healthy; MCI, mild cognitive impairment; ProD, prodromal; SCD, subjective cognitive decline; SEM, standard error of the mean.

## 3. RESULTS

### 3.1 Participant characteristics

A total of 287 participants were included in this study and categorized in each clinical diagnosis group as follows: CH; n=56, SCD; n=115, MCI; n=85, and AD; n=31 (**Table 1**). All participants reached approximately the same age (mean age >70 years old). Nevertheless, CH and SCD group were significantly younger than MCI and AD, as anticipated from a volunteer-based cohort. Sex was equally balanced between men and women except for CH and SCD groups where more females were recruited. Not surprisingly, *APOE* ε4 carriers were more frequent in AD and MCI groups compared to CH and SCD.

Clinical and sociodemographic data as well as blood assessment for all participants in each clinical diagnosis group is also presented in **Table 1**. No significant difference was found concerning the clinical and sociodemographic data which included nutritional scale, education and general health condition (waist size, weight, BMI, cardiac rhythm). Concerning blood assessment, we obtained normal range levels for cell count, glycemia index, vitamins and hormones, inflammation marker, renal and hepatic functions and lipid profile. No difference between clinical groups were observed except for renal function markers, with both urea nitrogen and creatinine levels being higher in AD group. This difference was no longer significant when age and sex were considered by calculating the glomerular filtration rate. Taken together, these results are consistent with the biological effect of age and sex on renal function rather than a true renal dysfunction associated with AD. As biomarker analyses were already adjusted for age and sex, no additional correction for renal function was performed. Group comparisons for cognitive and neuropsychiatric clinical scores, CSF biomarkers and standardized MRI brain volumes are presented in **Table 2** [36]. Overall, MCI and AD groups showed lower cognitive performance and more pronounced mood symptoms than CH and SCD, while SCD also presented with higher anxiety levels than CH. As expected, lower CSF Aβ42 and Aβ42/Aβ40 levels were found in AD participants compared to other clinical groups; however, this analysis was limited by the small subgroup with available CSF data (n=60). Likewise, lower global and regional brain volumes were observed with more advanced clinical stages.

**Table 2.**
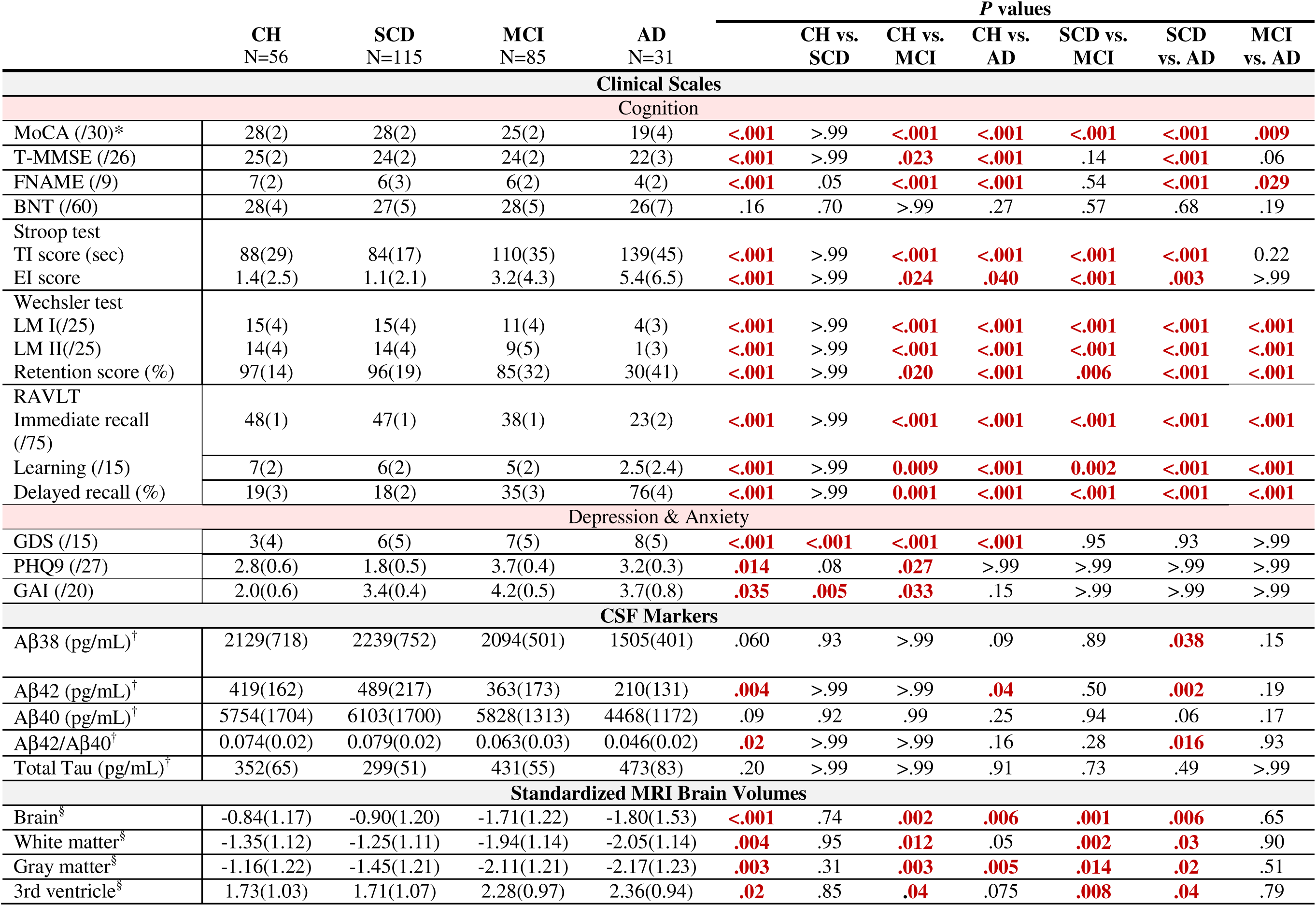
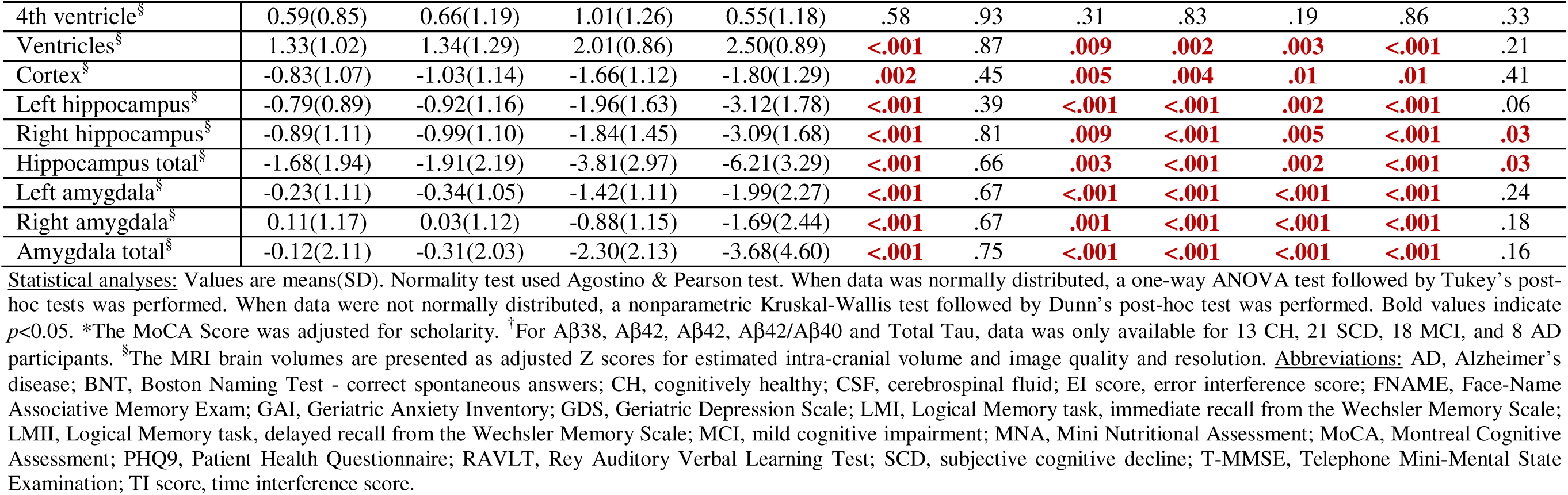
Clinical Scales, CSF Markers and Standardized MRI Brain Volumes in Each Clinical Diagnosis Group.

Information regarding the use of medications that could influence metabolic markers is provided in the **Supplemental Material Appendix D**. Overall, 119 participants (41.5%) were receiving lipid-lowering therapy, predominantly statins, whereas only 17 participants (5.9%) with diabetes were taking hypoglycemic agents. None of the participants were on basal insulin treatment, and only one (0.3%) was taking mealtime rapid-acting insulin.

### 3.2 Influence of demographic factors on peripheral metabolic markers across clinical groups

Both adiponectin (**Figure 1A,B**) and FGF-21 (**Figure 1E,F**) serum levels did not differ between clinical groups. Regarding IGFBP-2, a significant difference was found between CH and AD/MCI groups (**Figure 1I**). This difference remained significant even after adjustment for age and sex for AD versus CH (**Figure 1J**). Interestingly, the comparison between CH and SCD became significant after adjustment, raising the possibility that increased IGFBP-2 levels may represent an early event in the clinical progression, potentially associated with future risk of conversion to AD (**Figure 1J**). Among the three metabolic markers, only FGF-21 and IGFBP-2 showed a positive correlation with age (**Figure 1G,K**). Not surprisingly, we found higher levels of adiponectin in women compared to men regardless of clinical diagnosis, except in AD where this difference was not statistically significant (**Figure 1D**). No sex effect was observed for FGF-21 and IGFBP-2 (**Figure 1 H,L**).

### 3.3 Dysmetabolism is associated with lower levels of adiponectin and IGFBP-2, but higher levels of FGF-21

As expected, adiponectin, IGFBP-2 and FGF-21 were strongly correlated with determinants for metabolic syndrome and related disorders (see heatmap in **Figure 2A**). While levels of adiponectin and IGFBP-2 decreased along the rise of signs of metabolic defects, we observed a completely opposite effect for FGF-21, which was positively associated with the same markers. The strongest correlations included waist size, weight, BMI (**Figure 2B-D**), glycemia index (**Figure 2E-G**) as well as lipid profile data (triglycerides, high-density lipoproteins; **Figure 2H-M**). Similar results were obtained after age and sex adjustments. Therefore, although FGF-21 is recognized for its beneficial effects on metabolic health, we report here a paradoxical increase of this hormone in response to metabolic disturbances. The contrasting behaviors of these peripheral markers under metabolic stress make them valuable biomarkers for investigating the connections between metabolic dysfunction and AD, both in terms of their involvement and their specific behavior in this pathological context.

### 3.4 Higher levels of FGF-21 and IGFBP-2 are associated with impaired cognitive performance

If AD was only the central expression of a peripheral metabolic problem, the same pattern of associations as in dysmetabolism would be expected with cognitive performance. However, both FGF-21 and IGFBP-2 tended to be more elevated with impaired cognitive function and higher mental health burden, while adiponectin did not correlate with any of the clinical scales (**Figure 3**). Indeed, we found multiple significant correlations between FGF-21 or IGFBP-2 and cognitive scores, but only those between TMT part A and RAVLT immediate score and FGF-21 (**Figures 3F,I**) as well as between IGFBP-2 and MoCA (**Figure 3D**) remained statistically significant after age and sex adjustments. In other words, results show an association between lower performance in attention, working memory and processing speed subdomains and higher serum levels of FGF-21 and an association between lower global cognition and higher levels of IGFBP-2. The positive correlation between IGFBP-2 and GDS (**Figure 3M**) also suggested an indirect association between mood symptoms and cognitive decline. Thus, while our results show decreased peripheral levels of IGFBP-2 with metabolic dysfunction, its levels rather increase in the context of AD-associated cognitive decline.

### 3.5 Association between peripheral metabolic markers and AD biomarkers in the blood

Reciprocal correlations between peripheral metabolic markers and their associations with AD fluid-based biomarkers are presented in **Figure 4**. As expected, we observed highly significant reciprocal correlations between serum FGF-21, IGFBP-2 and adiponectin. More precisely, we found an inverse association between FGF-21 and adiponectin (**Figure 4B**) and a positive association between IGFBP-2 and adiponectin (**Figure 4C**), which remained significant even after age and sex adjustment. No significant correlation was found between adiponectin, FGF-21 or IGFBP-2 and all AD CSF biomarkers analyzed, that is; Aβ38, Aβ40, Aβ42, Aβ42/Aβ38, and total tau (**Figure 4A**). These results should be interpreted with caution, however, given the small number of participants with available CSF data, which may have limited the statistical power of the analyses. Finally, we also studied the relationship between metabolic markers and those of tau pathology in the plasma (pTau181, pTau231 and pTau217). We found that IGFBP-2 positively correlated with serum level of pTau181 (**Figure 4D**). A similar trend was found between IGFBP-2 and serum levels of pTau231 and pTau217, but this association did not reach statistical significance (**Figure 4A**). Hence, beyond its elevation observed in the context of cognitive decline, serum IGFBP-2 levels also appear to increase in association with plasma biomarkers indicative of neurodegeneration in AD.

### 3.6 Association between peripheral metabolic markers and regional brain volumes

Concerning MRI data, no association was found between adiponectin and all examined relative brain volumes (**Figure 5A**). FGF-21 levels were inversely associated with third ventricle and total ventricular volumes, a proxy of subcortical atrophy, after adjustment for age and sex (**Figure 5B**). Non-adjusted IGFBP-2 serum levels were found to be significantly inversely associated with total brain volume and several regional measurements including white matter, third ventricle, hippocampus and amygdala volumes. After adjustment for age and sex, only the correlation with the amygdala volume remained significant (**Figure 5C**). Nonetheless, these findings suggest that an elevation of serum IGFBP-2, rather than a decrease as seen in dysmetabolism, occurs during AD-associated neurodegeneration, even though this condition is itself linked to central insulin resistance.

### 3.7 Exploratory analysis of IGFBP-2 in prodromal AD

Because of the distinct association between IGFBP-2 and plasma pTau levels, we performed additional exploratory analyses in prodromal subjects. We defined a prodromal stage (ProD) in participants without a diagnosis of AD, but with an elevated blood level of pTau, in accordance with the current collective effort toward a biological definition of AD [43]. We chose plasma pTau instead of CSF because of insufficient number of subjects with available CSF sample. Similarly, tau PET was not included in the CIMA-Q protocol. The threshold for a biological or prodromal diagnosis was set at 8 pg/ml, which corresponds to one standard error of the mean lower than the mean of the three tau biomarkers (pTau181, pTau217, and pTau231) in subjects with an AD diagnosis.

Individuals who had a value of pTau181, pTau217, or pTau231 above 8 pg/ml were considered prodromal. This ProD group included 33 individuals from CH, SCD or MCI groups. Among these five clinical groups (CH, SCD, MCI, ProD and AD), with ProD individuals analyzed separately from their initial diagnostic categories, both AD and ProD individuals had the highest levels of IGFBP-2 (**Figure 6A**). After adjustment of IGFBP-2 for age and sex, the difference between CH and SCD was significant, whereas higher levels were detected in all cognitively impaired groups, but not ProD group compared to CH (**Figure 6B**). Despite the limited sufficient statistical power and the exploratory nature of these analyses, our observations may suggest an early elevation of circulating IGFBP-2 in AD, occurring independently of the prodromal stage as defined by a rise in pTau levels.

## 4. DISCUSSION

We explored the relationship between key peripheral markers of metabolism and AD-associated cognitive decline. We first established that adiponectin, FGF-21 and IGFBP-2 show distinct association with dysmetabolism and cognition. Whereas signs of metabolic disorders are associated with a rise in FGF-21, they are rather correlated with a decrease in adiponectin and IGFBP-2. We then show that both FGF-21 and IGFBP-2 increase along with age and lower cognitive performance. No association was found between adiponectin and cognition. Overall, IGFBP-2 is more strongly associated with clinical diagnosis, cognitive function and markers of neurodegeneration than FGF-21 including serum pTau and brain atrophy, and its increase may be an early event in the progression of cognitive decline in AD. The contrasting associations of FGF-21 and IGFBP-2 with cognitive disorders offer novel insights into how peripheral metabolic disturbances relate to AD.

### 4.1 Adiponectin, FGF-21 and IGFBP-2 in dysmetabolism

Very strong associations were observed between the three peripheral metabolic markers and metabolic dysfunction. Amongst them, clinical and biochemical evidence of obesity (higher waist size, weight and BMI), insulin resistance (elevated glycemia and glycated hemoglobin) and dyslipidemia (higher triglycerides and lower high-density lipoproteins) were associated with lower adiponectin and IGFBP-2, but higher FGF-21. This pattern of changes aligns with the extensive body of data accumulated over the years with adiponectin [9, 44, 45], IGFBP-2 [46–49] and FGF-21 [50–52]. For IGFBP-2, the observation that bariatric surgery reverses its obesity-induced decline is particularly compelling [53]. These divergent hormonal changes suggest distinct regulatory mechanisms of their production and release. See **Supplemental Material Appendix E** for more details regarding age and sex differences in peripheral metabolic markers.

### 4.2 Metabolic markers in relation to AD spectrum

Serum IGFBP-2 levels differed between groups across the AD spectrum, with SCD and most importantly AD participants presenting higher levels compared to controls. This observation is in accordance with previous reports indicating that a clinical diagnosis of AD is associated with higher concentrations of IGFBP-2 in the blood or CSF [25–27, 30–33, 35, 54, 55]. Recent proteomics data pinpoint IGFBP-2 as an early AD biomarker. Among a subset of 24 promising candidates selected from over 1000 blood proteins, IGFBP-2 was identified as being significantly associated with AD diagnosis, left entorhinal cortex atrophy and a faster rate of cognitive decline [32]. More recently, a comprehensive profiling study of plasma proteins in MCI participants revealed that IGFBP-2 was part of an 18-protein panel that could capture the profile changes of plasma proteome in MCI and AD [56]. However, these reports did not look at prodromal stages. Our finding of elevated IGFBP-2 in SCD compared to CH group is novel, yet coherent with existing literature. This may represent an early metabolic event contributing to the progression of age-related cognitive decline or AD.

Group comparisons did not reveal significant differences for FGF-21 or adiponectin. Only one published report showed an increased plasmatic level of FGF-21 in MCI compared to controls [57]. Findings on adiponectin are mixed, with some studies reporting reduced plasma, serum, or CSF levels in MCI and AD [18, 20, 21], while others show no difference [17, 19]. Thus, IGFBP-2, either measured on CSF or blood, stands out as having a greater potential as a biomarker for the early identification of individuals at higher risk of developing AD.

### 4.3 Metabolic markers in relation to cognition

Evidence linking peripheral metabolic markers to cognition remains limited. Here, IGFBP-2 showed the strongest association, with higher levels correlating with poorer global cognitive performance on MoCA. A similar trend was observed with FGF-21, although it did not reach significance. There was however a significant inverse correlation between FGF-21 and TMT part A completion time, a measure of attention and processing speed, and between FGF-21 and RAVLT Immediate score, a measure of working and episodic memory [58]. A few studies have reported an association between plasma IGFBP-2 concentrations and impaired episodic memory [26] and abstract reasoning [27]. One recent study show an inverse association between CSF IGFBP-2 and longitudinal changes in delayed memory and visuospatial abilities in an asymptomatic cohort of participants with a high genetic risk of AD, defined as a positive parental history [30]. However, the authors did not look at serum concentrations of IGFBP-2. Regarding FGF-21, at least two studies explored its association with cognition. One found the same inverse correlation with MoCA [59], while the other found no association with global cognition as assessed by Mini-Mental State Evaluation [50]. This may be explained by the higher sensitivity of MoCA for the detection of MCI [60].

Given the link between metabolic disorders and AD, the increase in FGF-21 in both conditions is expected. More surprising, however, is the opposite pattern observed with IGFBP-2. While AD is associated with brain insulin resistance [1, 6, 61], and peripheral insulin resistance typically correlates with lower IGFBP-2, we instead observed higher IGFBP-2 levels with AD progression. This inverse relationship, along with the association of elevated IGFBP-2 with age and cognitive decline, while being rather low in diabetes and obesity, provide new clues about the possible molecular mechanisms governing the role of IGFBP-2 in these diseases.

### 4.4 Metabolic markers and neurodegeneration

Midlife diabetes and obesity have been associated with an increased risk of cognitive decline [4], but not with neuropathology or imaging markers [8]. Thus, a strong association between metabolic hormones and CSF, blood or MRI endpoints was not expected. Still, we found a significant association between serum IGFBP-2 and plasma pTau181, a peripheral biomarker of AD pathology [38]. These associations were not significant with pTau217 and pTau231. Previous studies reported positive correlation between CSF-derived IGFBP-2 with both Aβ and total and phosphorylated tau CSF levels [24, 28, 30]. It has been proposed that nascent AD pathology may induce an upregulation in IGFBP-2 levels, possibly also explaining the positive association found between IGFBP-2 and Aβ levels [30, 62].

IGFBP-2 negatively correlated with hippocampus and amygdala volumes, suggesting a potential impact on AD-related atrophy [63]. This finding supports earlier reports linking higher plasma IGFBP-2 to reduced grey matter in AD-sensitive regions, such as the superior frontal and angular gyri [28, 64]. Higher plasma IGFBP-2 levels have also been associated with AD-specific brain atrophy patterns, indicated by smaller hippocampal volumes, among amyloid-negative individuals [26]. Measured in the CSF, IGFBP-2 is also negatively associated with entorhinal volume and cortical thickness in the piriform, temporal, and precuneus regions [30]. Taken together, these findings suggest that peripheral IGFBP-2 might be a better predictor of cognitive function and brain structural integrity, while CSF IGFBP-2 might more closely reflect AD-associated CSF amyloid and tau pathologies. Our findings further support an association between circulating IGFBP-2, but not FGF-21 or adiponectin, and AD neuropathology.

### 4.5 Cause or consequence?

IGFBP-2 has both central and peripheral origin. It is expressed by the hippocampus and the choroid plexus [16], but overall, most IGFBP-2 comes from the liver [65]. IGFBP-2 has been shown to cross the blood-brain barrier in a single study [66], but transport between the choroid plexus and CSF can also occur, and decreases with age [67]. Animal studies suggests that IGFBP-2 may play a role in insulin signaling resistance and altered neuronal energy metabolism in the hippocampus of AD brains [29, 68]. In mice, IGFBP-2 overexpression has been linked to resistance against age-related obesity, glucose intolerance, insulin resistance, and hypertension [69]. Paradoxically, while IGFBP-2 may potentially protect against dysmetabolism in older adults [69, 70], high levels also exhibit a negative effect on bone health and was found to be related to greater mortality [47, 48, 70, 71]. The role of IGFBP-2 remains unclear, as it may exert contrasting effects on peripheral metabolism and age-related disorders. A potential mechanistic link between IGFBP-2 and tau pathology in AD is that elevated IGFBP-2 may reduce IGF signalling by sequestering IGFs, thereby impairing IGF-I–mediated inhibition of tau phosphorylation and contributing to the formation of neurofibrillary tangles.

In humans, the bulk of FGF-21 is liver-derived [72] but it is also expressed in skeletal muscles and brain [73]. As stated above, previous studies repeatedly reported upregulation of FGF-21 in association with obesity, dyslipidemia and insulin resistance in the elderly [50–52]. It is believed that increased mitokines is an adaptive response to stress [50] and that FGF-21 synthesis and release may be a compensatory mechanism to restore metabolic control [74]. However, chronic metabolic conditions can eventually provoke FGF-21 resistance, increasing the individual’s sensitivity to metabolic disorders and accelerating the aging process [50, 75]. Thus, a plausible hypothesis that emerges is that FGF-21 represents a beneficial compensatory phenomenon for both metabolic syndrome and the prevention of AD.

### 4.6 Study limitations

This study was conducted using a large, well-characterized cohort of 287 participants classified into four clinical groups, including SCD. To our knowledge, no previous study has examined and compared adiponectin, FGF-21 or IGFBP-2 levels in this emerging preclinical stage. Beyond its focus on early AD stages, the study’s novelty lies in its combined analysis of three insulin signaling pathway molecules and its comprehensive clinico-biochemical assessment. This represents the first detailed and integrative analysis of metabolic biomarkers in the early phases of AD. Nevertheless, some limitations must be noted. The unbalanced group sizes, especially the relatively small AD group, may have limited the statistical power of the comparison analyses. The cross-sectional design precludes conclusions about causality, and follow-up data (2–6 years) are still being collected. Longitudinal analyses may clarify the role of these markers in disease progression. The predominantly Caucasian, Quebec-based, and highly educated volunteer sample also limits generalizability, and a validation cohort may be necessary to ensure reproducibility. Lastly, metabolites were measured in serum, while IGFBP-2 in CSF may also relate to AD pathology [28, 30]. Thus, future studies should investigate metabolic biomarkers in a wider range of body fluids, including CSF.

## Abbreviations

Aβ: amyloid beta;
AD: Alzheimer’s disease;
BMI: body mass index;
BNT: Boston Naming Test;
CH: cognitively healthy;
CIMA-Q: Consortium for the early identification of Alzheimer’s Disease-Quebec;
CSF: cerebrospinal fluid; ELISA, enzyme-linked immunosorbent assays;
FGF-21: fibroblast growth factor 21;
FLAIR: fluid-attenuated inversion recovery;
FNAME: Face-Name Association;
GAI: Geriatric Anxiety Inventory;
GDS: Geriatric Depression Scale;
IGF: insulin-like growth factor;
IGFBP-2: insulin-like growth factor-binding protein 2;
MCI: mild cognitive impairment;
MoCA: Montreal Cognitive Assessment;
MRI: magnetic resonance imaging;
PET: positron emission tomography;
PHQ: Patient Health Questionnaire;
ProD: prodromal stage;
RAVLT: Rey Auditory Verbal Learning Test;
SCD: subjective cognitive decline;
SEM: standard error of the mean;
tMMSE: Telephone Mini-Mental State Examination;
TMT: Trail Making Test;
WDSST: WAIS Digit Symbol Substitution Test.

## ACKOWLEDGEMENTS / CONFLICTS / FUNDING SOURCES / CONSENT STATEMENT

## Data Availability

All data produced in the present study are available upon reasonable request to the authors.

## Acknowledgements

C.D.T. interpreted the results and wrote the manuscript. H.L.D. analyzed the data, interpreted the results and contributed to redaction. R.C. analyzed the data and contributed to figures. C.T. performed adiponectin, FGF-21, IGFBP-2 and pTau217 ELISA assays and formatted results. A.P. contributed to data analysis and creation of graphs and figures and participated to the redaction. J.V.E. and M.L. participated in the initial data analysis of FGF-21 and J.V.E. also contributed to redaction.

A.L. did half of FGF-21 ELISA assays. M.T. conceptualized the study, participated in the ethics committee and grant applications, contributed to data acquisition and revised the manuscript. O.P. provided brain MRI data and manuscript revision for important intellectual content regarding neuropsychological testing. S.B. contributed to initial funding. A.G. revised the final manuscript with a focus on clinical and metabolic implications and contributes to the CIMAQ cohort by identifying healthy CSF for control.

F.P. contributed to initial funding and revised the final manuscript. H.Z. generated pTau231 and pTau181 data. F.C. conceptualized the study, supervised the study process, and revised the manuscript for important intellectual content. We gratefully thank the participants in the Consortium for the Early Identification of Alzheimer’s Disease – Quebec (CIMA-Q) for their contribution.

## Consent Statement

All human subjects included in this study provided written informed consent.

