## Supplemental Material for "Distinct alterations of adiponectin, FGF-21 and IGFBP-2 link dysmetabolism with cognitive decline across the Alzheimer’s disease spectrum"

**Appendix A**

**Inclusion criteria for all participants:**

1. Be aged 65 or older
2. Live in a community or a residence for an independent person (or an equivalent environment)
3. Have a score ≥ 17 on the telephone-mini mental state examination (T-MMSE)
4. Understand, read, and write French or English
5. Have sufficient visual and auditory acuity to be able to go through the neuropsychology tests visit
6. Be accompanied during clinical visits for participants with mild Alzheimer’s Disease. Have an informant to answer questions in person, by phone, or in writing for other participants.
7. Be willing to: answer questionnaires about one’s state of health; have a physical and neuropsychological evaluation; submit to a blood test.

**Exclusion criteria for all participants:**

1. Plan to move out of Quebec in the next 3 years
2. Have a score ≥ 20 on the Patient Health Questionnaire-9 (PHQ-9) scale
3. Have a score > 1 on the Clinical dementia rating (CDR)
4. Have a disease or impairment of the central nervous system (including subdural hematoma (active or past), subarachnoid hemorrhage (active or past), primary or metastatic brain cancer, epilepsy (active), dementia (other than mild Alzheimer’s Disease) or other neurodegenerative diseases).
5. Have had intracranial surgery
6. Have an active addiction to alcohol, drugs, or narcotics
7. Have a regular consumption of benzodiazepines greater than the equivalent of 1mg/day lorazepam taken orally
8. Have an illness/condition that is related to cognitive impairment or one that could interfere with the participant’s participation in the project

**Table 1. Inclusion Criteria Test Results for Each Clinical Diagnosis Group**

| **Clinical Diagnosis Group** | **Jessen** | **MoCA (/30)** | **Wechsler** | **CDR** | **NIA-AA**  **Clinical Criteria** |
| --- | --- | --- | --- | --- | --- |
| **CH** | A or B | ≥ 26 | I. ≥ 9 for 16+ years of education II. ≥ 5 for 8-15 years of education III. ≥ 3 for 0-7 years of education | 0 |  |
| **SCD** | C | ≥ 26 | I. ≥ 9 for 16+ years of education II. ≥ 5 for 8-15 years of education III. ≥ 3 for 0-7 years of education | 0 |  |
| **Early MCI** | B or C | 20-26 | I. 9-11 for 16+ years of education II. 5-9 for 8-15 years of education III. 3-6 for 0-7 years of education | 0.5 | Fit for MCI-AD |
| **Late MCI** | B or C | 20-25 | I. ≤ 8 for 16+ years of education II. ≤ 4 for 8-15 years of education III. ≤ 2 for 0-7 years of education | 0.5 | Fit for MCI-AD |
| **AD** | B or C | 13-25 | I. ≤ 8 for 16+ years of education II. ≤ 4 for 8-15 years of education III. ≤ 2 for 0-7 years of education | (0.5) 1 | Fit for probable Alzheimer |

Abbreviations: CH, cognitively healthy; SCD, subjective cognitive decline; MCI, mild cognitive impairment; AD, Alzheimer’s disease; MoCA, Montreal Cognitive Assessment; CDR, Clinical Dementia Rating; NIA-AAA, National Institute on Aging and Alzheimer’s Association.

**Appendix B**

**Table 2. Routine Blood Analysis**

Complete blood count

International Normalized Ratio (INR)

Thyroid Stimulating Hormone (TSH)

Sodium, potassium, and calcium

**Blood glucose:**

Fasting glycaemia

Glycosylated hemoglobin (HA1C)

Blood Urea Nitrogen (BUN)

Vitamin B_12_ and Vitamin B_9_

**Liver function tests:**

Aspartate transaminase (AST)

Alanine transaminase (ALT)

Alkaline phosphatase (ALP)

C-Reactive Protein

**Blood lipids:**

Total Cholesterol (Chol)

Low-Density Lipoprotein-Cholesterol (LDL-Chol)

High-Density Lipoprotein-Cholesterol (HDL-Chol)

Triglycerides (TG)

**Appendix C**

**Appendix D**

**Supplementary Table 1.** Use of lipid-lowering and antidiabetics medication among CIMA-Q participants

| **Medication** | **Number of participants (%)** |
| --- | --- |
| **Lipid-lowering drugs** | **119 (41.5%)** |
| Statins | 116 (40.4%) |
| Cholesterol absorption inhibitors | 4 (1.4%) |
| Fibrates | 4 (1.4%) |
| **Antidiabetics** | **17 (5.9%)** |
| Biguanides | 14 (4.9%) |
| Sulfonylureas | 3 (1.0%) |
| DPP-4 inhibitors | 4 (1.4%) |
| SGLT2 inhibitors | 2 (0.7%) |
| GLP-1 agonists | 1 (0.3%) |
| Basal insulin | 0 (0%) |
| Mealtime insulin | 1 (0.3%) |

Medication use was confirmed by the pharmacy, either through a medication list provided by the patient from their pharmacy or through a verification call made by the evaluating physician. Abbreviations: DPP-4, dipeptidyl peptidase-4; SGLT2, sodium-glucose transport 2; GLP-1, glucagon-like peptide-1.

**Appendix E**

**Discussion on peripheral metabolic markers and their association with age and sex**

In agreement with the present findings, previous studies have shown a strong correlations between FGF-21 [1, 2], IGBFP-2 [3] and age. It has been hypothesized that the increase in FGF-21could be a response to latent age-related diseases or to low-level inflammation and cellular stresses [4]. While higher levels of IGFBP-2 in older individuals have been reported [5-8], this association does not appear to be linear. The serum concentration of IGFBP-2 first decreases between birth and puberty, followed by a steady increase, especially after 60 years [5, 9]. In a longitudinal cohort, the age-related increase in IGFBP-2 remained significant even after controlling for sex, BMI, insulin-like growth factor 1 levels and insulin sensitivity, suggesting its presence as a normal expected finding with aging, independently of dysmetabolism [3]. In sum, our results support the idea that advancing age is associated with increased secretion of FGF-21 and IGFBP-2 by the liver, but whether it is beneficial or detrimental remains unknown.

The women recruited in the CIMAQ cohort exhibited higher blood levels of adiponectin, as reported in other populations [10, 11]. Several previous studies have reported changes in adiponectin levels in women between pre-, peri-, and post-menopause stages, as female sex steroid hormones, primarily estrogens, influence adipogenesis and adipose tissue metabolism [12-14]. These sex-specific metabolic processes may affect adiponectin levels in older adults. However, our data contrast with reports of significant sex differences in FGF-21 levels, which show higher levels in young adult men (21-59) and the oldest-old women (82-99) [15]. These sex-specific differences may not have been captured by our more age-restricted sample. Finally, the absence of sex differences in IGFBP-2 levels is consistent with findings from another study [16]. Overall, the associations found between FGF-21 or IGFBP-2 and age, metabolism, and cognition in the present study were not significantly influenced by differences in biological sex.

**Related references**

**Appendix F**

**Results adjusted for multiple comparisons (Benjamini-Hochberg)**

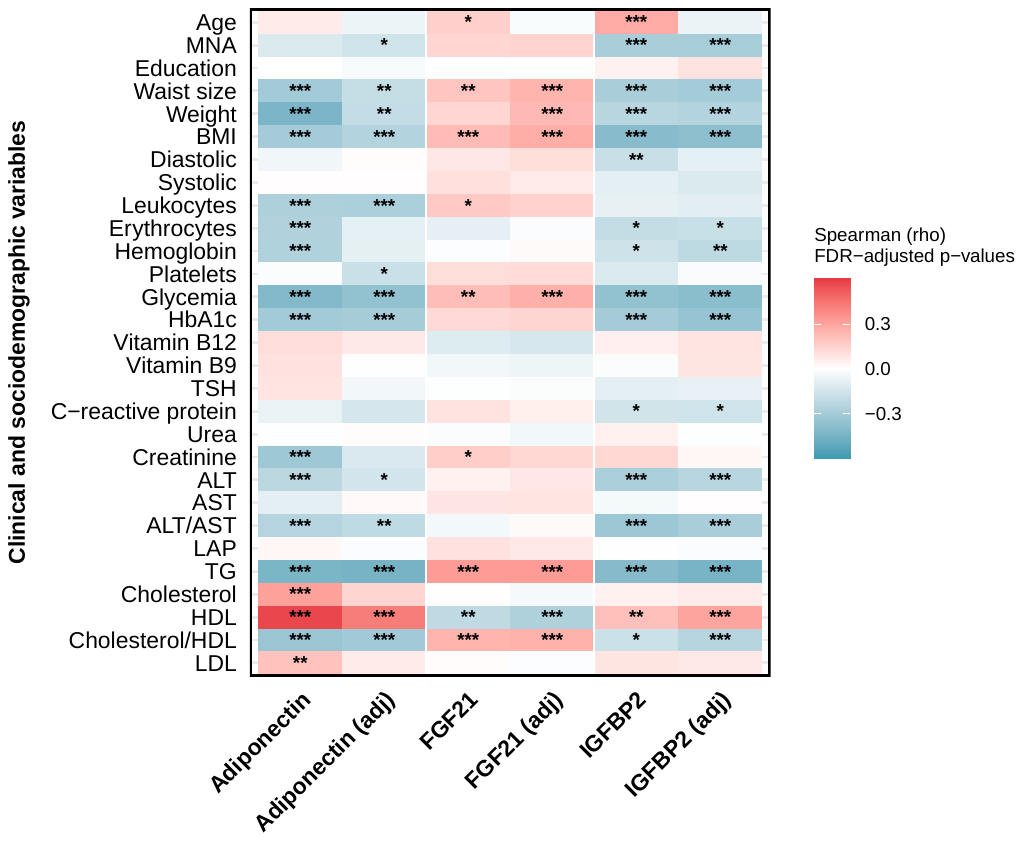

**Supplementary Figure 1. Multiple linear regression for Adiponectin, FGF-21 and IGFBP-2 on Clinical and Sociodemographic Data & Blood Assessment.** Statistical analyses: Red and blue highlighted cells respectively indicate significant positive and negative correlations. We present both unadjusted and adjusted p-values (for age and sex) from Spearman correlation analyses. A Benjamini-Hochberg correction was applied to account for multiple comparisons, and significance levels are displayed as follows: *p < 0.05, **p < 0.01, ***p < 0.001, ****p < 0.0001**.**

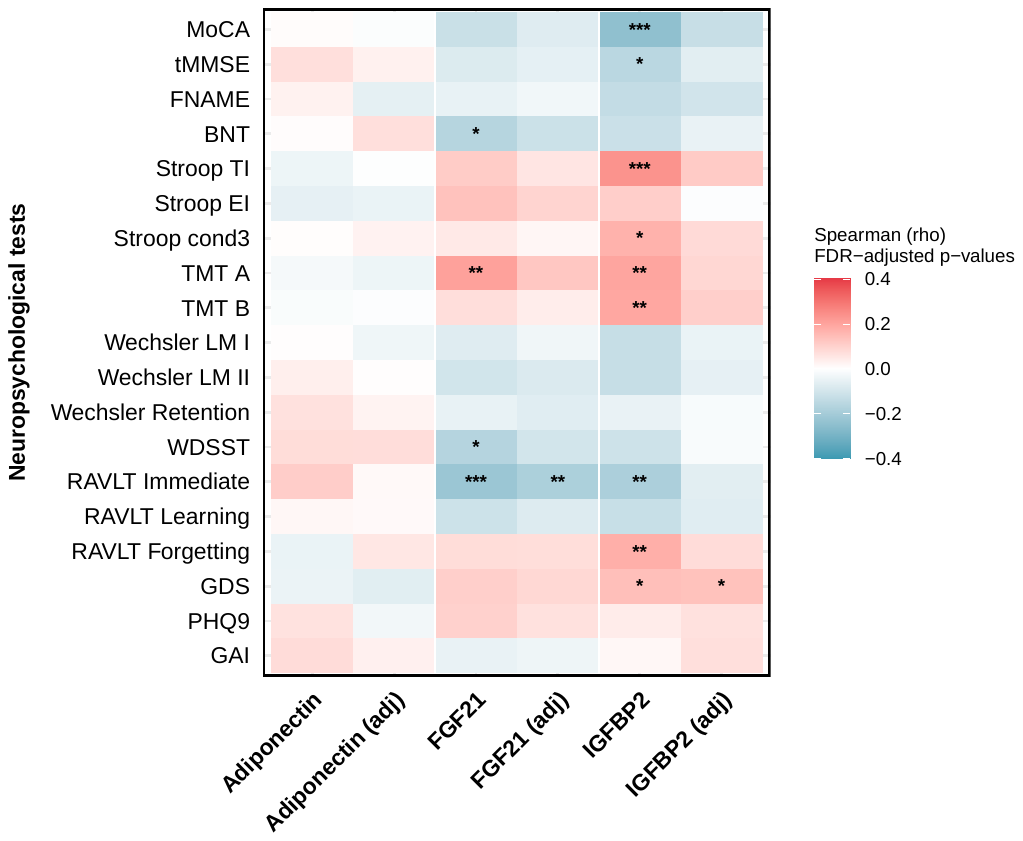

**Supplementary Figure 2. Multiple linear regression for Adiponectin, FGF-21 and IGFBP-2 on Clinical scores.** Statistical analyses: Red and blue highlighted cells respectively indicate significant positive and negative correlations. We present both unadjusted and adjusted p-values (for age and sex) from Spearman correlation analyses. A Benjamini-Hochberg correction was applied to account for multiple comparisons, and significance levels are displayed as follows: *p < 0.05, **p < 0.01, ***p < 0.001, ****p < 0.0001**.**

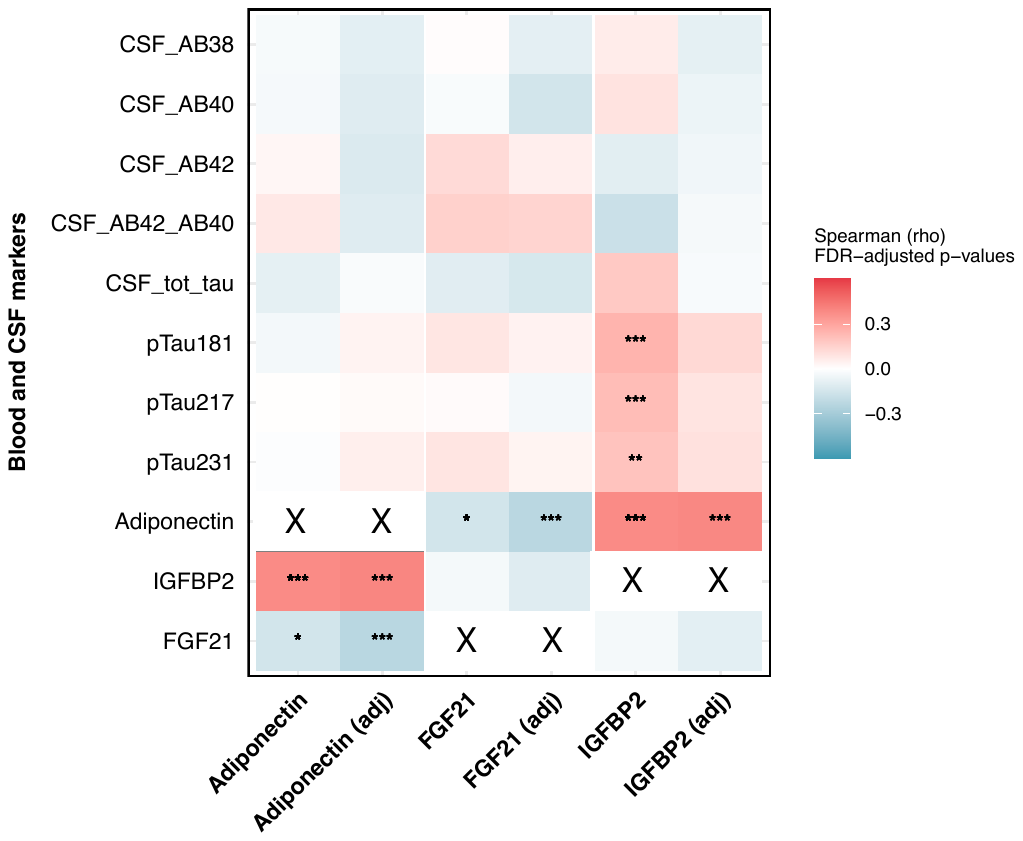

**Supplementary Figure 3. Multiple linear regression for Adiponectin, FGF-21 and IGFBP-2 on CSF markers and serum pTau.** Statistical analyses: Red and blue highlighted cells respectively indicate significant positive and negative correlations. We present both unadjusted and adjusted p-values (for age and sex) from Spearman correlation analyses. A Benjamini-Hochberg correction was applied to account for multiple comparisons, and significance levels are displayed as follows: *p < 0.05, **p < 0.01, ***p < 0.001, ****p < 0.0001**.**

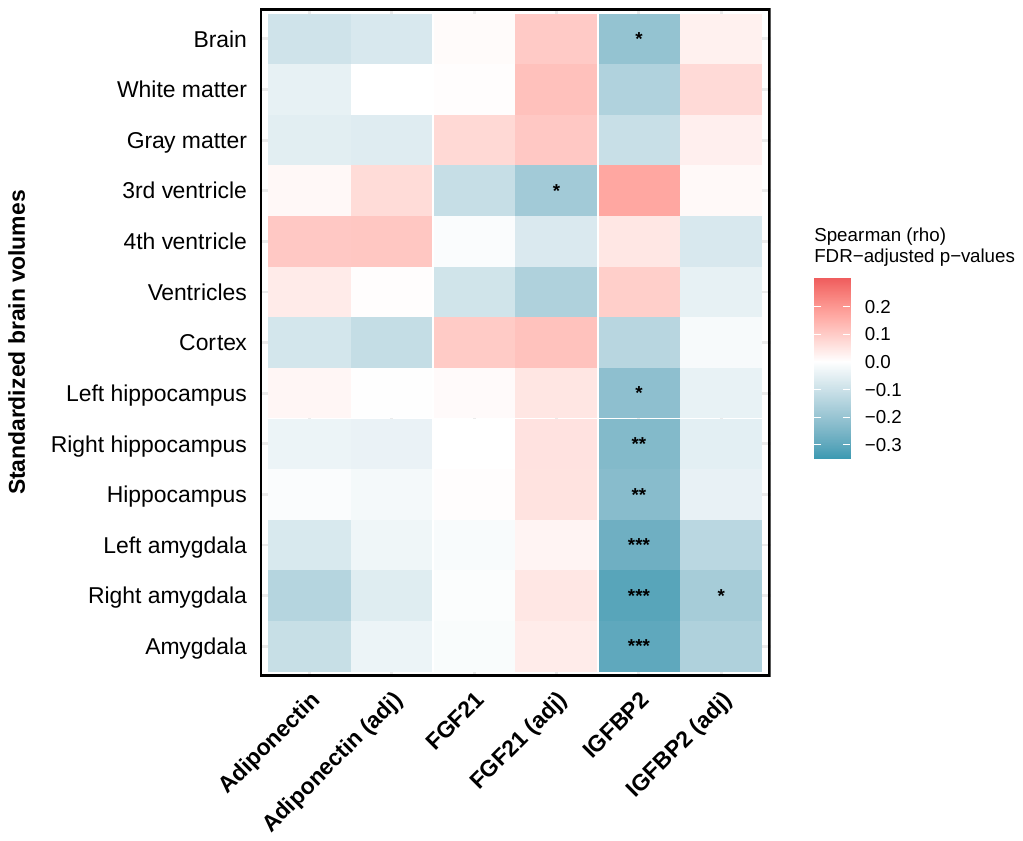
**Supplementary Figure 4. Multiple linear regression for Adiponectin, FGF-21 and IGFBP-2 on brain relative volume.** Statistical analyses: Red and blue highlighted cells respectively indicate significant positive and negative correlations. We present both unadjusted and adjusted p-values (for age and sex) from Spearman correlation analyses. A Benjamini-Hochberg correction was applied to account for multiple comparisons, and significance levels are displayed as follows: *p < 0.05, **p < 0.01, ***p < 0.001, ****p < 0.0001**.**
